# Development of an original algorithm to characterize serological antibody response that improve infectious diseases surveillance

**DOI:** 10.64898/2026.04.16.26350925

**Authors:** Solohery Lalaina Razafimahatratra, Lova Tsikiniaina Rasoloharimanana, Tokilalaniaina Mananjo Andriamaro, Paulo Ranaivomanana, Matthieu Schoenhals

## Abstract

Interpreting serological data remains challenging, particularly in low-prevalence or cross-reactive contexts, where antibody responses often show substantial overlap between exposed and unexposed individuals and may depart from normal distributional assumptions. Conventional cutoff-based approaches often yield inconsistent or biased estimates of seroprevalence. Here, we present a decisional framework based on finite mixture models (FMMs) that enhances the robustness and interpretability of serological analyses. Beyond simply applying mixture models, our framework integrates multiple methodological innovations : (i) systematic comparison of Gaussian and skew-normal mixture models to accommodate asymmetric antibody distributions; (ii) rigorous model selection using the Cramér–von Mises test (p > 0.01) combined with a parsimonious score (APS) to prioritize models with well-separated clusters; and (iii) hierarchical clustering of posterior probabilities to collapse latent components into biologically meaningful seronegative and seropositive groups.

Applied to chikungunya virus (CHIKV) data from Bangladesh, the framework produced prevalence estimates consistent with ROC-based methods while probabilistically identifying borderline cases. Validation on SARS-CoV-2 and dengue datasets further demonstrated its generalizability: for SARS-CoV-2, the approach identified up to five latent clusters with high sensitivity (up to 100%) and specificity (up to 100%), enabling discrimination by disease severity. For dengue, it revealed interpretable subgrouping consistent with background exposure and subclinical infection, despite limited confirmed cases.

By integrating distributional flexibility, robust goodness-of-fit testing, and biologically guided cluster consolidation, this decisional FMM framework provides a reproducible and scalable method for serological interpretation across pathogens and epidemiological settings, addressing key limitations of threshold-based classification.

## Introduction

Serological data play a central role in epidemiology and public health. By quantifying antibody responses to pathogens, serology provides critical insights into infection prevalence, immune protection, and vaccine-induced responses. These data are essential not only for infection surveillance but also for guiding public health interventions and evaluating immunization programs. However, the classification of individuals as seropositive or seronegative remains a challenge. Antibody distributions often overlap due to heterogeneous exposure histories and cross-reactive immune responses, making it difficult to establish clear thresholds that distinguish exposed from unexposed populations. This ambiguity can lead to misclassification, bias in prevalence estimates, and reduced sensitivity in monitoring population-level immunity.

Several existing and widely used techniques for threshold definition have been applied in serological studies. One simple and widely applied approach is the mean of presumed negative controls plus three standard deviations (mean + 3SD), which is easy to implement but highly dependent on the representativeness of the negative samples and sensitive to outliers^1^. Another common method is receiver operating characteristic (ROC) curve analysis, which offers a statistically robust way to balance sensitivity and specificity when gold-standard positive and negative samples are available; however, its utility is limited when such reference materials are lacking^2^. Finally, two-component finite mixture models attempt to disentangle overlapping antibody distributions by assuming that populations are composed of seronegative and seropositive subgroups ^3^. This approach avoids the need for gold-standard samples and provides a flexible framework, but it relies heavily on model assumptions and may oversimplify reality, as additional modes in the distribution can carry biological meaning, for example, reflecting antibody cross-reactivity, waning immunity in certain individuals, or signals of recent exposure, suggesting that serological responses should not always be forced into a strict binary classification. While these approaches provide practical solutions, each has inherent limitations, and none fully resolve the ambiguity in threshold determination, underscoring the continued need for methodological refinement in serological studies.

Finite mixture models (FMM) have emerged as a powerful statistical tool to address these challenges. Unlike fixed cut-off approaches, mixture models account for heterogeneity in serological responses by representing the observed data as a combination of distinct underlying subpopulations (e.g., seronegative vs. seropositive groups). This probabilistic framework is particularly suited to serological data, which often exhibit skewness, variability, and overlapping distributions.

The objective of this article is to introduce a flexible finite mixture model framework for analyzing serological data. By incorporating Gaussian and skewed components, the proposed method aims to better capture the underlying heterogeneity of antibody responses. We also outline a validation strategy for model performance and discuss how cluster-based interpretation can inform the classification of seropositive and seronegative individuals. This framework provides a more robust and generalizable approach to serological data analysis, with the potential to improve accuracy in epidemiological studies and public health decision-making.

## Methodology

Analysing serological data from surveillance studies can be challenging when no predefined cutoff exists. Standard approaches rely on well-characterized samples, biologically confirmed positives (e.g., by PCR or culture), confirmed negatives, or presumed negatives such as samples from non-endemic areas or collected during the pre-pandemic period. In these cases, classical methods such as receiver operating characteristic (ROC) curves or the mean plus three standard deviations (Mean + 3 SD) can be used to define a threshold for classifying positive samples. However, when such well-characterized samples are unavailable, alternative strategies are required. To address this, we developed a novel algorithm based on finite mixture models (FMM), which consider the overall sample distribution as a combination of one or more components. FMMs are widely used in serology, but our approach incorporates a decision-making algorithm and parametric metrics to guide the selection of the optimal number of components representing the population.

First, the variance of the data was reduced by applying logarithmic and square-root transformations, providing both strong (log) and moderate (sqrt) variance stabilization. Each transformed dataset was then fitted using both Gaussian and skewed mixture models, reflecting common practices in serological analyses. The selection of the best model and transformation was performed using the Cramér–von Mises goodness-of-fit test (p-value). Among models showing acceptable fit, the parsimonious score (PS) was used to identify the model with the lowest complexity. Standard model selection metrics such as BIC or AIC were not used here, as we needed to compare models across different transformations. In cases where multiple models (GMM and SMM) had equivalent PS values, the effective sample size (n_eff) was used as an additional metric to assess the robustness of each component. The model with the highest n_eff was considered the most reliable. Once the optimal model was identified, the number of components (k) was evaluated. When k≤2, the population could be characterized as seronegative and seropositive, or seronegative only. For k>2, unsupervised hierarchical clustering of the components was applied to group them into two main clusters, enabling a binary classification of the population (Figure 1).

**Figure 1:**
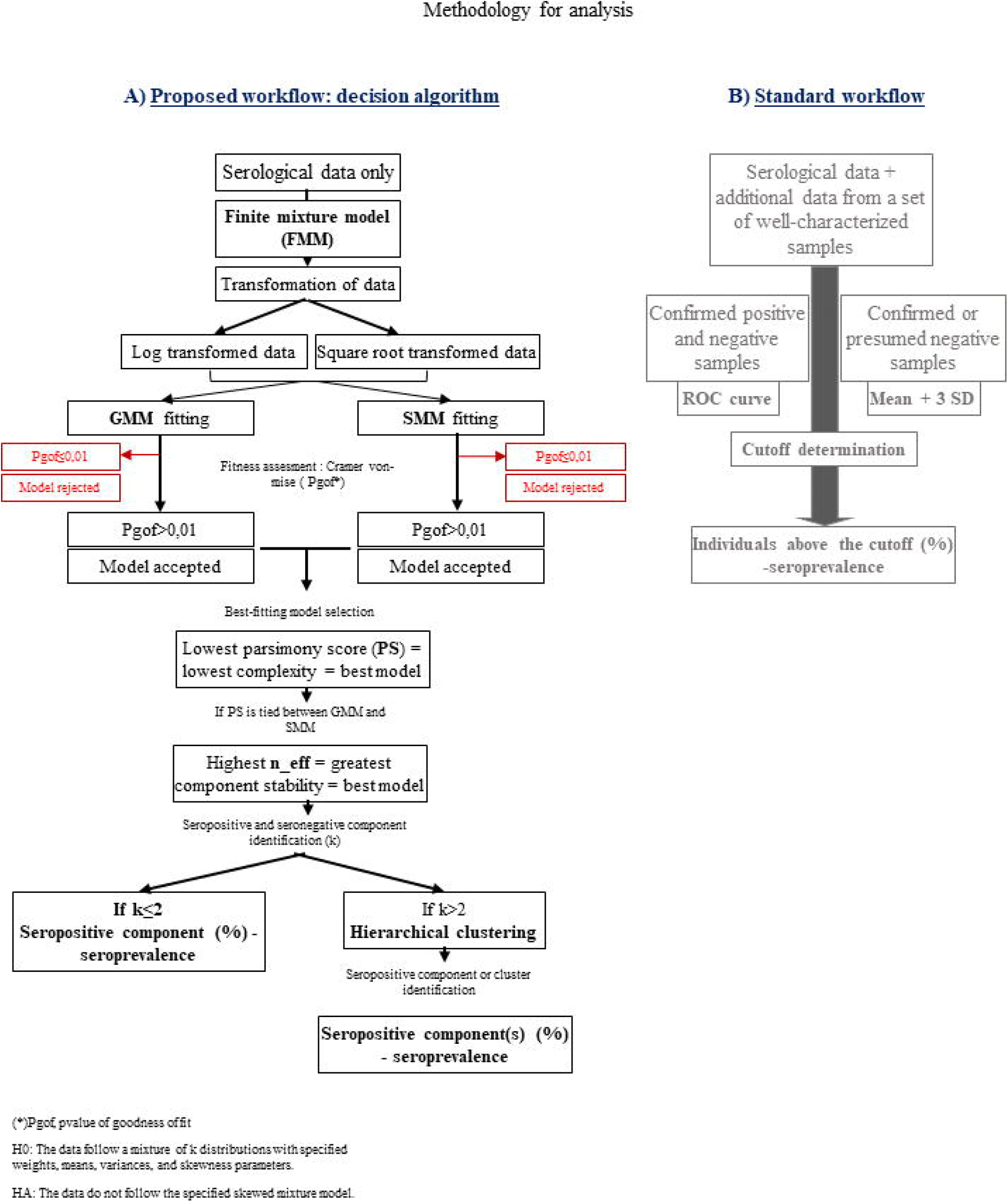
Workflows for serological data analysis: standard and decisional approaches. (A) Decisional algorithm workflow. (B) Standard workflow. FMM, finite mixture model; GMM, Gaussian mixture model; SMM, skewed mixture model; n_eff, effective sample size; ROC, receiver operating characteristic; SD, standard deviation; k, number of component.

The decisional algorithm was validated using serological datasets from three independent studies, obtained either from publicly available data repositories or directly from the original authors. Across these datasets, the algorithm consistently enabled the classification of seronegative and seropositive populations. The following sections describe the methodological framework underlying the decisional algorithm, including the finite mixture models employed, model selection criteria based on goodness-of-fit p-values and parsimony scores, stability assessment using effective sample size (n _eff_), and the hierarchical clustering strategy used to consolidate mixture components into biologically meaningful groups.

### Overview of Finite Mixture Models

We modeled the distribution of serological measurements using finite mixture models (FMMs), which assume that the observed data arise from a mixture of *K* latent subpopulations, each described by a probability density function *f*_*k*_ (*y* | θ_*k*_)^4,5^. The overall density is given by :

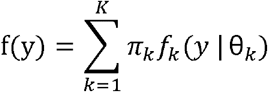

where *πk* is the mixing proportion for component 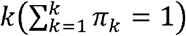 and *θ*_*k*_ represents the set of component-specific parameters.

In this analysis, we considered two types of components: Gaussian and skew-normal. Gaussian components are parameterized by location (*μ*_*k*_) and 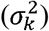, representing the mean and variance of the distribution :

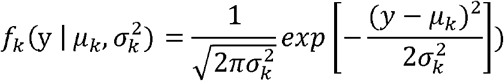

Skew-normal components extend the Gaussian by including a skewness parameter (*α_k_*) to model asymmetric distributions often observed in serological data^6^. The skew-normal density is defined as :

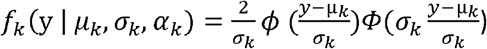

Where *ϕ* (·) and *Φ* (·) are the standard normal density and cumulative distribution functions, respectively. Here, μ_*k*_ and *σ*_*k*_ control the location and scale, while *σ_k_* determines the degree and direction of skewness.

Parameter estimation was performed using maximum likelihood via the Expectation– Maximization (EM) algorithm, which iteratively optimizes the likelihood of the observed data given the mixture model. At each iteration, the EM algorithm computes the expected component membership for each observation (E-step) and then updates the parameters (*π*_*k*_, *μ*_*k*_, *σ*_*k*_, *α*_*k*_) to maximize the likelihood (M-step). This procedure continues until convergence, producing parameter estimates that best explain the observed data in terms of the mixture model.

### Model Selection and Algorithm

The selection of the optimal model was based on a combination of statistical adequacy, parsimony, and stability^6^.

The adequacy of each candidate model was first evaluated using the Cramér–von Mises test statistic, which measures the squared distance between the empirical distribution function *F*_*n*_(*y*) and the fitted cumulative distribution function 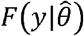:

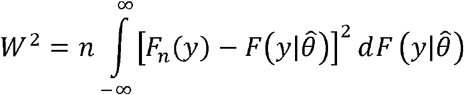

Where *n* is the sample size and 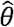 denotes the set of estimated parameters. The corresponding *p*-value (*P*_*gof*_) was obtained by parametric bootstrap. A model was considered to provide an adequate description of the data when *P*_*gof*_ >0.01, indicating that the null hypothesis of correct specification could not be rejected. Models failing this criterion were discarded from further consideration.

Among models satisfying the goodness-of-fit requirement, parsimony was evaluated using the parsimony adjusted score (APS), which balances model fit and complexity :

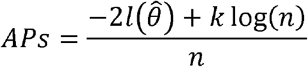

Where 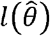 is the maximized log-likelihood, *k* is the number of free parameters, and *n* is the sample size. This criterion is conceptually related to the Bayesian Information Criterion (BIC) but is normalized by the sample size, facilitating comparability across datasets of varying sizes. Lower APS values indicate models that achieve a better trade-off between fit and simplicity^7^.

In addition, we examined the effective sample size of each fitted component to assess the stability of the parameter estimates. For a mixture component *k*, the effective sample size was defined as :

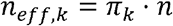

Where *π*_*k*_ is the estimated mixing proportion for component *k* and *n* is the total sample size. Components with an effective sample size *n*_*eff,k*_<10 were considered weakly supported and therefore potentially unstable. This threshold was not intended as a strict statistical cutoff but as a pragmatic criterion to avoid over-interpretation of components driven by very small numbers of observations, for which parameter estimates are known to be highly sensitive to sampling variability in mixture models. Similar minimum-size considerations are commonly adopted in mixture modeling to ensure numerical stability and interpretability of component-specific estimates. Importantly, this criterion was applied in conjunction with goodness-of-fit and parsimony measures, and not in isolation. In scenarios where a small component reflects a biologically plausible subgroup (e.g., a rare seronegative population in a highly prevalent setting), such components can be retained provided that the overall model remains well supported and stable.

The final decision process therefore proceeded in three stages. First, models failing the goodness-of-fit criterion were rejected. Second, among the models retained, the ones with the lowest APS were selected. Third, in cases where two or more models showed comparable APS values, the model with the largest minimum effective sample size across components was chosen. The number of mixture components was then consolidated using hierarchical clustering in order to ensure biological interpretability. When more than two components were identified, they were grouped into two biologically meaningful categories, corresponding to seronegative and seropositive populations, while allowing for additional substructure to reflect heterogeneity in immune response.

The clustering procedure was based on the posterior probabilities of belonging to a component membership^8,9^. For each individual *i* and component *j*, the posterior probability is given by:

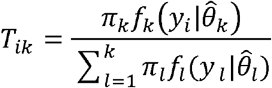

Where *π*_*k*_ is the mixing proportion, 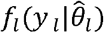 is the density of component *k* evaluated at *y*_*i*_, and 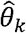 are the estimated parameters of component *k*

To quantify the similarity between components, we first computed the average posterior probability profiles for each component, defined as :

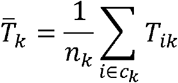

where *n*_*k*_ is the number of individuals assigned to component *k*, and 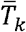 is the mean posterior probability vector for that component. These vectors summarize the typical assignment profile of each mixture component.

Pairwise distances between components were then calculated using a correlation-based dissimilarity measure:

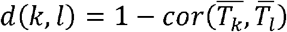

where 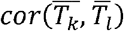 is the Pearson correlation between the average posterior profiles of components *j* and *l*. This formulation ensures that components with highly similar assignment structures are considered close in the clustering space.

The dissimilarity matrix was used as input for agglomerative hierarchical clustering with the average linkage method. At each step of the algorithm, the distance between two clusters *A* and *B* was defined as :

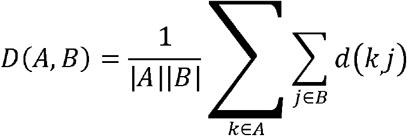

Where |*A*| and |*B*| are the number of components in clusters *A* and *B*, respectively. The dendrogram provided a hierarchical representation of similarity among the *k* mixture components. For interpretability, the tree was cut to yield two main groups, broadly corresponding to seronegative and seropositive populations. The mapping between the original mixture components and the two main groups was established by minimizing within-group distances while preserving the original mixing proportions of the model.

The robustness of this hierarchical clustering strategy lies in its ability to aggregate mixture components in a data-driven, non-parametric way. By focusing on correlation of posterior profiles rather than raw parameter values, the method captures the biological similarity of components while reducing over-fragmentation. This ensures that the final classification retains sensitivity to heterogeneous serological response patterns while yielding a parsimonious and interpretable grouping.

### Evaluation of Sensitivity and Specificity

For the finite mixture model (FMM), sensitivity and specificity were estimated by cross-tabulating the model-based cluster assignments against well-characterized reference classifications. Specifically, the cluster containing the highest proportion of reference-positive individuals was designated as “positive,” while all other clusters were treated as “negative.” A confusion matrix was then constructed, from which sensitivity and specificity were calculated:

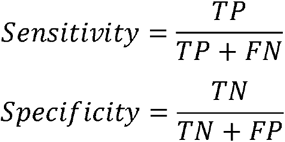

where *TP* denotes true positives (patients correctly classified as positive), *FN* false negatives (patients misclassified as negative), *TN* true negatives (healthy individuals correctly classified as negative), and *FP* false positives (healthy individuals misclassified as positive).

### Validation of the model

#### Study 1 : Chikungunya serological data of Allen SW et al. 2024

One of the datasets used to validate the decisional algorithm originated from a nationwide serological study conducted in Bangladesh prior to the 2017 chikungunya virus (CHIKV) epidemic ^10^. In that study, the original authors aimed to assess prior CHIKV transmission and population immunity. A total of 2,938 individuals were sampled across 70 communities, with blood samples and questionnaire data collected. CHIKV-specific antibodies were measured using a multiplex immunoassay, which was calibrated against plaque reduction neutralization tests (PRNT). To define seropositivity, PRNT was performed on a subset of 89 samples, and the corresponding immunoassay measurements were used to construct a receiver operating characteristic (ROC) curve. Based on this analysis, the authors selected a fluorescence intensity threshold of 5.5 units, which maximized sensitivity and specificity and was subsequently used to estimate a seroprevalence of 2.4%, indicating minimal prior CHIKV circulation in the population.

The serological measurements from this study, kindly provided by Dr. Henrik Salje, one of the study’s authors (Department of Genetics, University of Cambridge, UK), were reanalyzed here using the proposed decisional algorithm. Using square root–transformed data, the algorithm selected a three-component mixture model, in which two components showed substantial overlap and were subsequently merged using hierarchical clustering (Figure 2). The resulting seropositive group corresponded to an estimated seroprevalence of 2.6%. When compared against the seropositivity classification defined by the ROC-based threshold reported in the original study, the decisional algorithm identified nine discordant samples classified as seropositive by the model but seronegative under the ROC-based definition. Using the published ROC-based classification as the reference standard, the algorithm achieved a sensitivity of 100% and a specificity of 99%.

**Figure 2:**
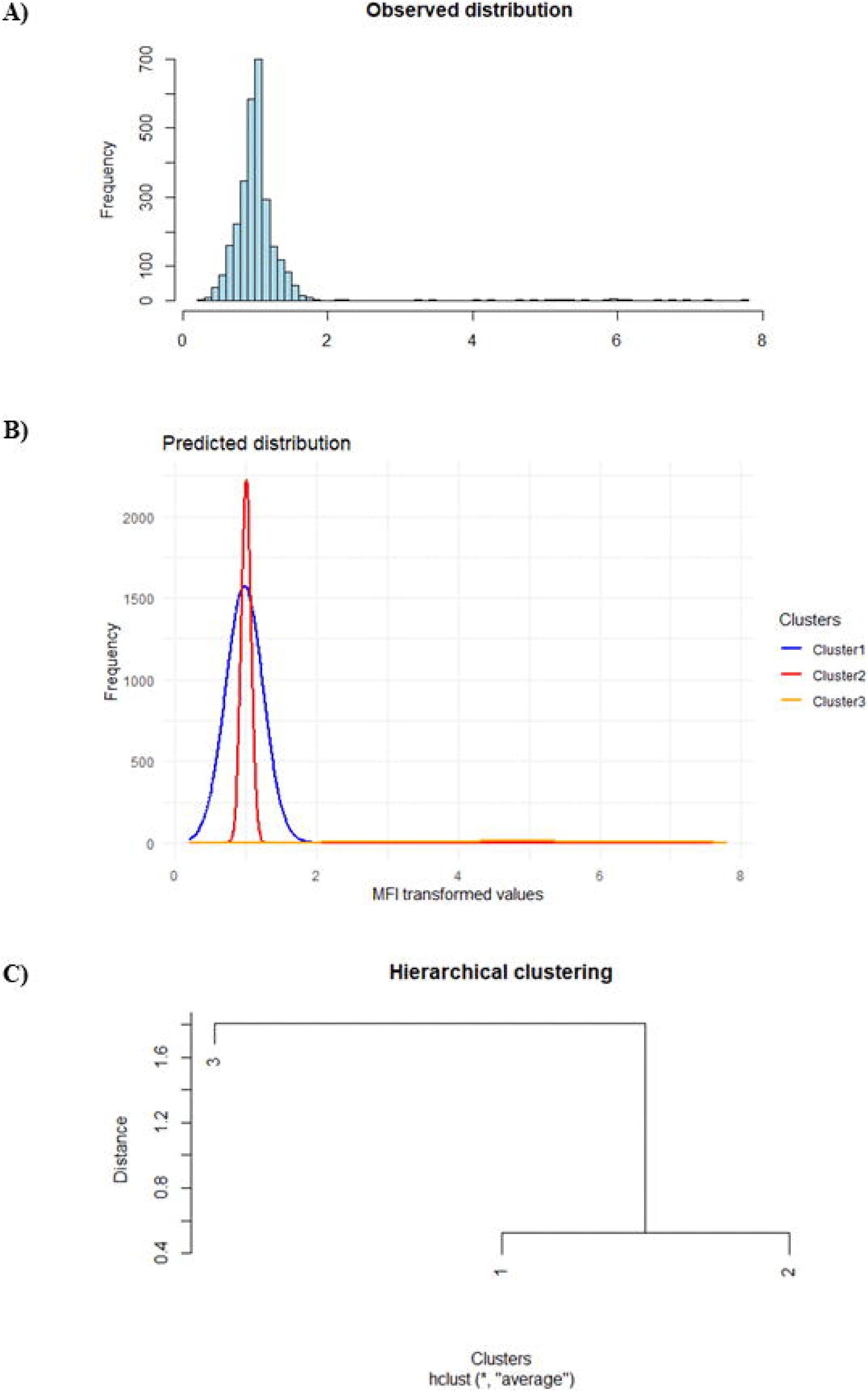
Distributions and clustering of serological reactivity to Chikungunya virus. (A) Observed distribution of population reactivity frequencies based on square-root transformed MFI data, (B) Model-predicted distribution of population reactivity using a skewed mixture model, (C) Hierarchical clustering of serological reactivity profiles

#### Study 2 : SARS-COV2 serological data of Yates et al. 2021

The original study investigated the relationship between SARS-CoV-2–specific antibody responses, particularly IgG subclass distributions and Fc receptor–related features, and COVID-19 disease severity ^11^. The dataset comprised 630 serum samples, including 94 prepandemic healthy controls and 536 convalescent, RT-PCR–confirmed COVID-19 cases collected at the Wadsworth Center (New York State Department of Health). Disease severity was self-reported and classified as mild (n = 217), moderate (n = 215), severe (n = 49), or uncharacterized (n = 55). Serological measurements were performed using a Luminex-based multiplex assay quantifying IgA, IgM, and IgG subclasses (IgG1–4) targeting the nucleocapsid (N), receptor-binding domain (RBD), and S1 and S2 regions of the spike protein. Antibody responses were expressed as median fluorescence intensity (MFI). In the original analysis, seropositivity thresholds were defined for each antigen–isotype combination as the mean plus three standard deviations (mean + 3 SD) of the prepandemic control samples.

The serological measurements from this study, obtained from the publicly available ImmPort repository^12^, were reanalyzed here using the proposed decisional algorithm. Of the 28 antigen–isotype variables available, 14 did not yield an acceptable fit under the mixture-model framework and were therefore excluded from the decisional analysis. The remaining 14 variables, for which adequate model fit was achieved, were retained and analyzed further. Across these datasets, the decisional algorithm identified between two and five latent mixture components, which were subsequently evaluated and consolidated according to the algorithmic decision rules. The results obtained from this reanalysis were then compared with the seropositivity classifications reported in the original study. Across antibody classes, the sensitivity of the FMM-based algorithm ranged from 17.8% (IgM_N) to 100% (IgG_S2), while specificity ranged from 70.4% (IgA_N) to 100% (IgG1_RBD and IgG1_S2). Under the conventional mean + 3 SD threshold, sensitivity varied from 13.1% (IgM_S2) to 99.4% (IgG_S2), and specificity from 96.8% (IgM_N and IgM_S2) to 98.9% (IgG3_S1, IgG1_S2, IgG3_S2) (Figure 3).

**Figure 3:**
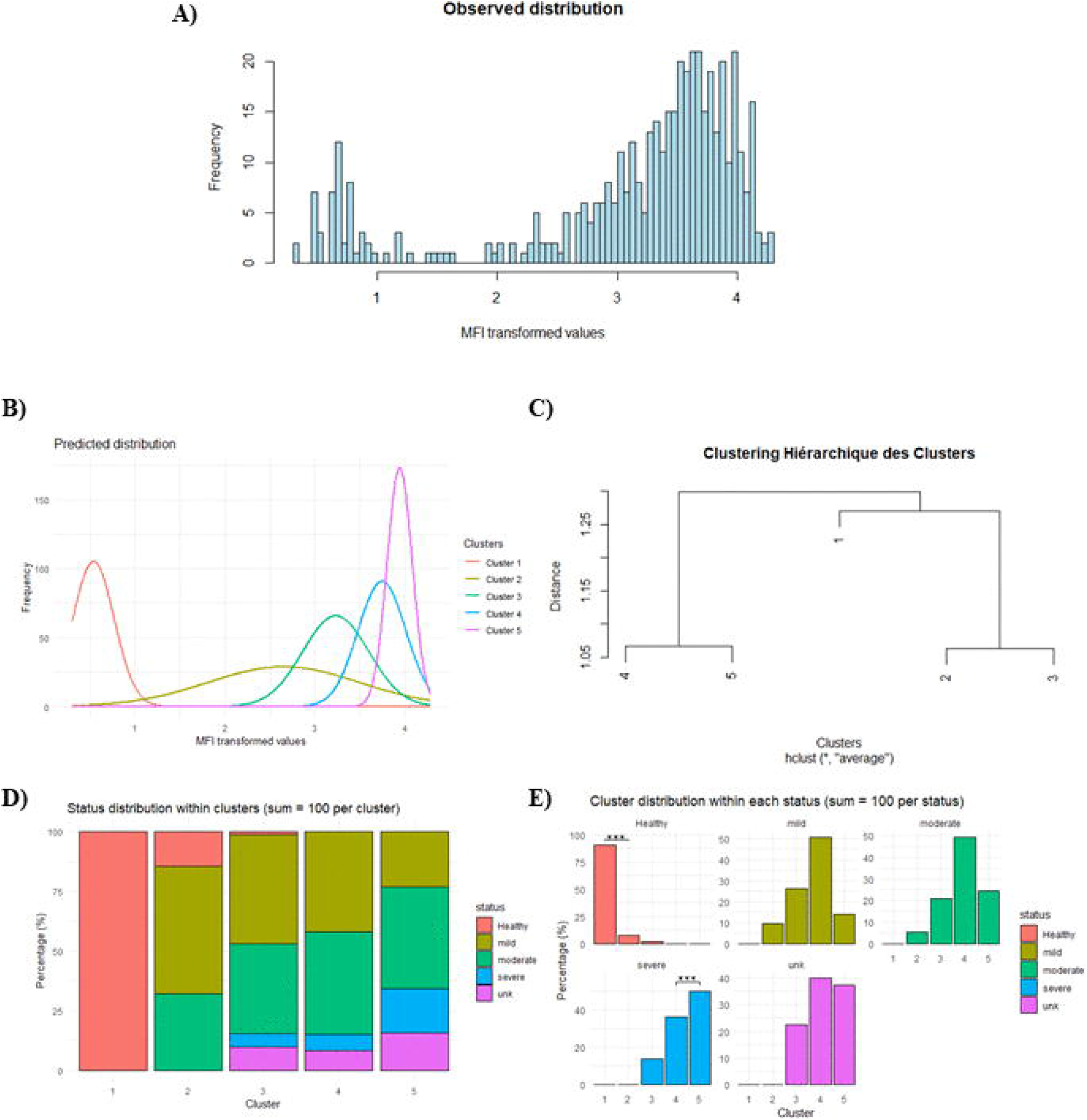
Performance comparison: mean + 3 SD threshold vs. decisional algorithm. (A) Mean + 3 SD threshold. (B) Decisional algorithm. N, nucleocapsid; RBD, receptor binding domain; S1 and S2, spike regions.

Balanced accuracy (BA), calculated as the average of sensitivity and specificity for each antibody 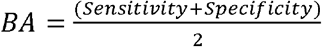, was used to summarize the overall diagnostic performance. The FMM algorithm yielded the highest BA for IgG3_S2 (97.7%), followed by IgG_S2 (95.3%) and IgG1_S1 (97.2%), indicating excellent discrimination between positive and negative sera. When averaged across all antibodies, the mean balanced accuracy was 86.5% for the FMM algorithm and 87.4% for the mean + 3 SD method. The mean sensitivity reached 79.1% for the FMM-based approach versus 71.8% for the mean + 3 SD method, while the mean specificity was 90.1% and 97.9%, respectively (Table 1).

**Table 1:** Comparative performance metrics of the conventional mean + 3 SD threshold and the decisional algorithm framework. Statistical significance was assessed using the Wilcoxon signed-rank test, with p < 0.05 considered significant.

|  | Mean + 3 SD | Decisional algorithm | p-value |
| --- | --- | --- | --- |
|  | 71.8 | 79.1 | 0.17 |
| Mean sensitivity (%) |  |  |  |
| Mean specificity (%) | 97.9 | 90.1 | **0.02** |
| BA_mean (%) | 87.4 | 86.5 | 0.85 |

BA, Balanced accuracy ; SD, standard deviation ; p-value < 0.05, statistically significant difference

Statistical comparisons between the standard threshold-based method and the decisional algorithm were performed using paired Wilcoxon signed-rank tests, with pairing defined at the level of individual serological variables (n = 14 variables). For each antigen, sensitivity, specificity, and balanced accuracy were computed under both methods, yielding paired performance measures for direct within-antigen comparison. The Wilcoxon signed-rank test was selected as a nonparametric approach that does not rely on distributional assumptions and is appropriate for paired comparisons across a limited number of variables.

Using this framework, the FMM-based algorithm exhibited a significantly lower specificity compared with the mean + 3 SD threshold method (p = 0.02), while no significant difference in balanced accuracy was observed (p > 0.05). This result indicates that, although the decisional algorithm slightly reduces specificity, reflecting a modest increase in false-positive classifications, it preserves overall diagnostic balance between sensitivity and specificity. In practical terms, the algorithm improves detection of true positives without compromising overall classification performance across serological markers.

An interesting finding emerged from the IgG1 response to the RBD, where the model identified five distinct clusters. Using the decisional algorithm, the hierarchical clustering for IgG1_RBD revealed that clusters 1, 2, and 3 corresponded to seronegative individuals, whereas clusters 4 and 5 grouped seropositive ones (Figure 4). When analyzed by clinical status, the clusters clearly differentiated disease categories, as reflected by the sensitivity and specificity values reported in Table 2. Within this framework, the decisional algorithm accurately classified individuals by clinical severity, clearly distinguishing healthy donors, mild/moderate cases, and severe cases (Figure 4). The model showed excellent performance in identifying healthy individuals, with a sensitivity of 90.7% and perfect specificity (100%). For mild to moderate cases, it achieved a sensitivity of 80.7% and a specificity of 74.4%, indicating strong discrimination despite expected overlaps between these intermediate categories. Moreover, the algorithm effectively captured the dynamics of individuals who became seronegative over time, particularly those represented in clusters 2 and 3, and to a lesser extent in cluster 1.

**Figure 4:**
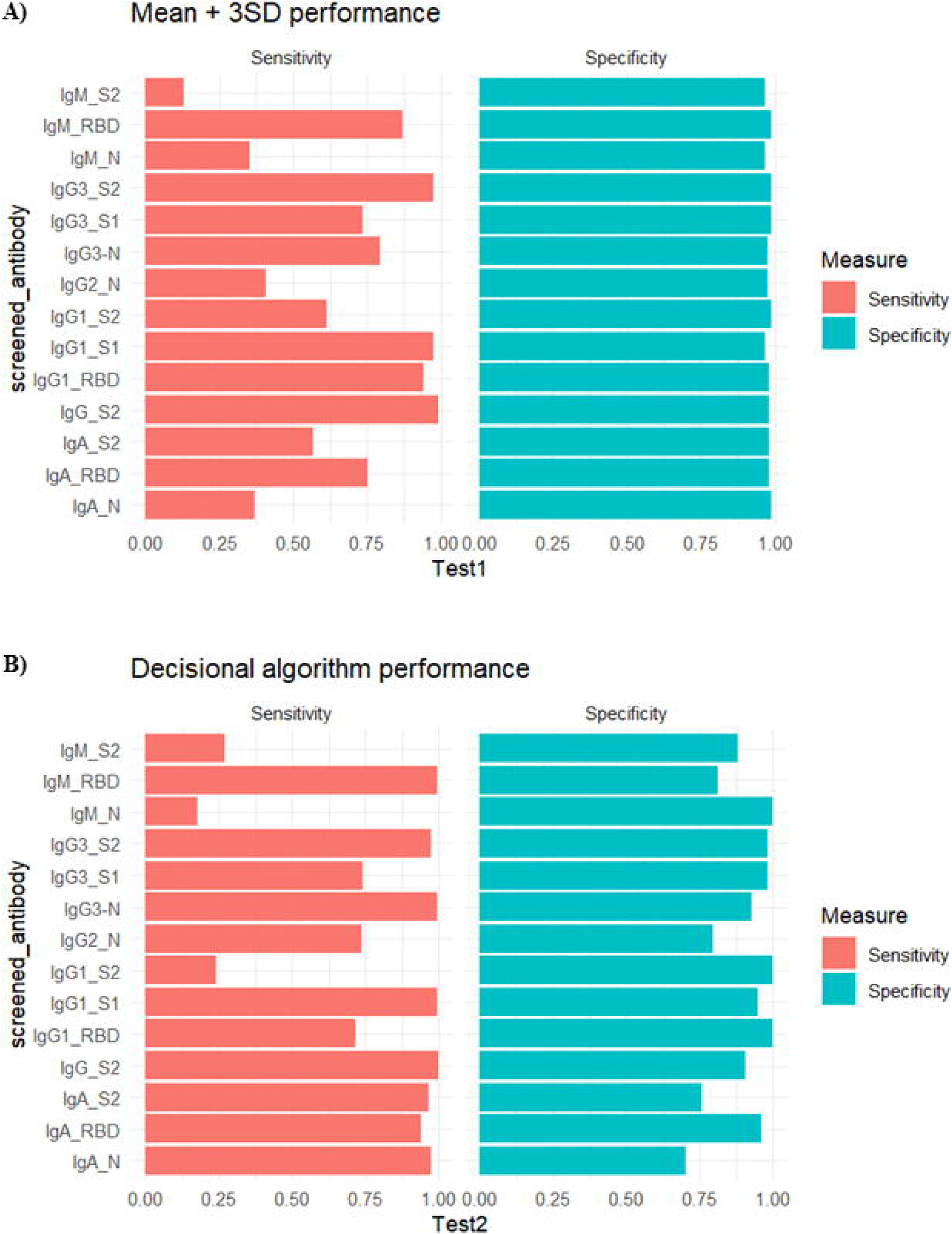
Model-predicted clusters for IgG1 anti-RBD and corresponding participant health status distribution. (A) Observed distribution of population reactivity frequencies based on square-root transformed MFI data, (B) Model-predicted distribution of population reactivity using a skewed mixture model, (C) Hierarchical clustering of serological reactivity profiles, (D) Status distribution within clusters, (E) Cluster distribution within each status.

**Table 2:**
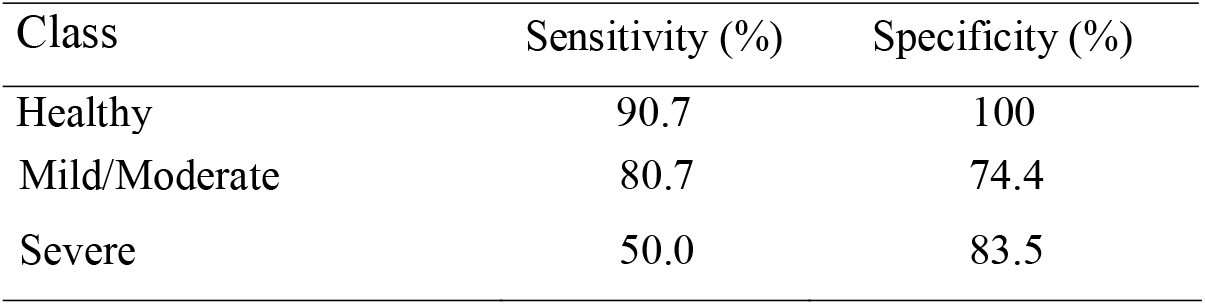
Classification performance of the decisional algorithm by clinical severity category (Healthy, Mild/Moderate, Severe).

| Class | Sensitivity (%) | Specificity (%) |
| --- | --- | --- |
| Healthy | 90.7 | 100 |
| Mild/Moderate | 80.7 | 74.4 |
| Severe | 50.0 | 83.5 |

#### Study 3 : Dengue serology data of Suárez-Medina et al. 2018

This investigation examined the relationship between prior dengue virus infection and markers of systemic inflammation in healthy three-year-old children in Havana, Cuba. A total of 865 children provided blood samples, of whom 14 (1.6%) had a documented prior clinical diagnosis of dengue infection, while 851 had no such diagnosis. Dengue-specific IgG levels were quantified using the Vircell assay and expressed as an antibody index. Because no validated serological cutoff exists for identifying prior dengue infection in this age group, the authors classified “elevated” IgG levels using a data-driven threshold defined as the median antibody index within the study population. Using this approach, they reported no significant difference in IgG titers between children with and without a prior clinical diagnosis of dengue, suggesting a high frequency of subclinical or unrecognized infections among children without documented disease.

The serological measurements from this investigation, obtained from the Figshare repository^13^, were reanalyzed here using the proposed decisional algorithm. After logarithmic transformation, the algorithm selected a four-component mixture model. Based on the resulting classification, the model differentiated individuals with and without a prior clinical dengue diagnosis with a sensitivity of 50% and a specificity of 60% (Figure 5). The modest sensitivity (50%) and specificity (60%) observed in this dataset should be interpreted in light of substantial limitations of the reference classification rather than as evidence of poor algorithmic performance. Prior dengue infection was defined based on parent-reported clinical diagnosis, which is known to substantially underestimate true dengue exposure in young children, given the high frequency of asymptomatic, mild, or misdiagnosed infections in this age group. Consistent with this limitation, the original analysis reported no significant difference in dengue IgG levels between children with and without a reported prior diagnosis, indicating weak discriminative signal in the serological measurements themselves. Under such conditions, any serology-based classification approach, whether threshold-based or model-based, is intrinsically constrained by label uncertainty and biological overlap between groups. In this context, the decisional algorithm identifies latent serological structure that is not fully captured by the clinical reference definition, highlighting the limitations of self-reported diagnosis as a gold standard rather than a failure of the modeling approach.

**Figure 5:**
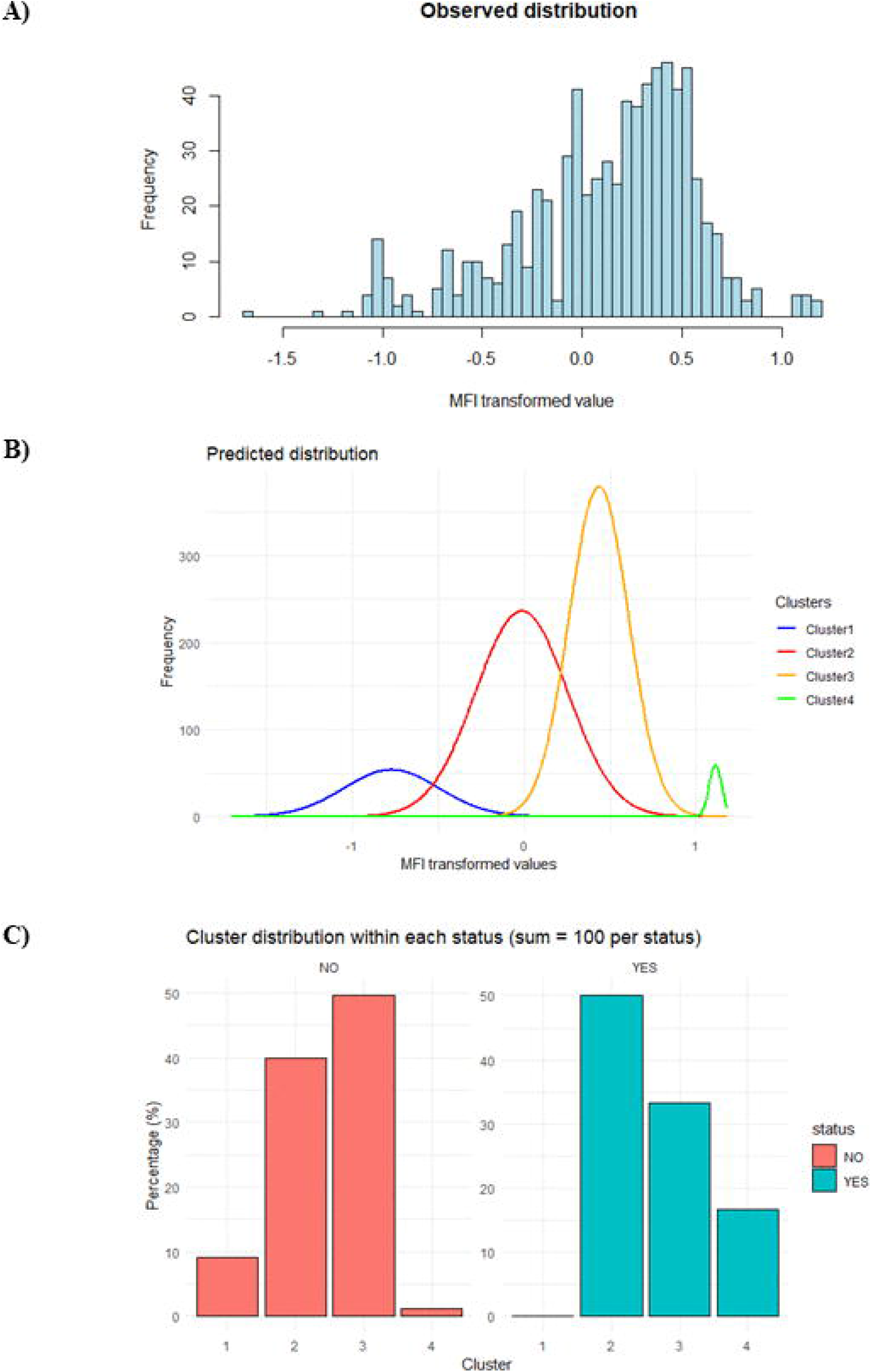
Model-predicted clusters for Dengue and corresponding participant status distribution. (A) Observed distribution of population reactivity frequencies based on square-root transformed MFI data, (B) Model-predicted distribution of population reactivity using a skewed mixture model, (C) Cluster distribution within each status. NO, no prior dengue (n=14); YES, prior dengue (n=851)

## Discussion

A key contribution of our study lies not simply in applying finite mixture models to serological data, but in the decisional framework we developed to guide interpretation of results. While FMMs have been previously used to estimate seroprevalence in the absence of reference standards^14,15^, most studies rely on a single model type, typically Gaussian mixtures, and interpret results directly from the best-fit solution. By contrast, we systematically compared both Gaussian mixture models (GMMs) and skew-normal mixture models (SMMs), allowing us to better accommodate the asymmetry often present in antibody distributions. To ensure robust model selection, we incorporated a multi-layered evaluation strategy. First, goodness of fit was assessed using the Cramér–von Mises test, a more stringent criterion than visual inspection or information criteria alone, with only models achieving p > 0.01 considered acceptable. Second, in cases where multiple models fitted adequately, we introduced a parsimonious score (APS) to prioritize models with the clearest separation of clusters, thus reducing the risk of overfitting and spurious components. Third, we extended interpretation by applying hierarchical clustering to the posterior probability profiles of latent clusters. This allowed us to collapse multiple subgroups into two biologically meaningful categories, seronegative and seropositive, while still preserving information about heterogeneity.

Application to a chikungunya virus (CHIKV) dataset from Bangladesh further illustrates the framework’s utility in low-prevalence settings. In such contexts, antibody distributions often exhibit strong overlap between negative and positive populations, making cutoff-based classification particularly prone to misclassification errors ^16,17^. By probabilistically identifying borderline cases, our approach achieved nearly identical prevalence estimates. This feature is especially relevant for arboviral infections such as CHIKV, where asymptomatic or mildly symptomatic infections are common ^18,19^ and where low pre-epidemic seroprevalence has been documented in other Asian populations prior to explosive outbreaks ^20,21^.

The framework also proved valuable in the context of SARS-CoV-2 serology. The algorithm identified between two and five latent clusters across antibodies, achieving high sensitivity (up to 100% for IgG_S2) and specificity (up to 100% for IgG1_RBD, IgM_N, and IgG1_S2). Balanced accuracy reached 97.7% for IgG3_S2, highlighting strong overall classification performance. When compared to the conventional mean + 3 SD threshold, the mean specificity was significantly higher for the mean + 3 SD method (98%) than for the FMM-based algorithm (90%) (p = 0.02), indicating that the conventional threshold was more conservative in correctly identifying true negatives. However, the balanced accuracy did not differ significantly between the two approaches, suggesting that the higher specificity of the mean + 3 SD method was counterbalanced by its lower sensitivity. Overall, the FMM-based algorithm maintained comparable diagnostic equilibrium while improving the detection of true positives. Moreover, the five-cluster solution for IgG1_RBD enabled meaningful stratification by disease severity, clearly separating severe cases and healthy controls, and partially distinguishing mild and moderate cases. These findings highlight the framework’s ability not only to improve sensitivity but also to extract clinically relevant heterogeneity from serological data.

In the dengue dataset, apparent classification performance was modest (sensitivity ≈50%, specificity ≈60%); however, this outcome primarily reflects limitations of the reference standard rather than deficiencies of the modeling framework. Prior dengue infection was defined by parent-reported clinical diagnosis, a measure known to substantially underestimate true exposure in young children due to the high prevalence of asymptomatic, mild, or unrecognized infections. Consistent with this limitation, the original study reported no significant differences in dengue IgG levels between children with and without a reported diagnosis, indicating limited discriminative signal in the serological data itself. In this context, the decisional algorithm nonetheless identified interpretable latent subgroupings that are consistent with background exposure and potential subclinical transmission. These results underscore the ability of the framework to extract biologically meaningful structure even in settings characterized by sparse, noisy, or imperfect gold standards.

Interpreting serological data presents several well-recognized challenges that limit the reliability of conventional cutoff-based approaches. In low-prevalence settings, antibody distributions are often skewed and overlapping, making it difficult to distinguish seronegative from seropositive individuals, as illustrated by Rogier et al. (2015) in Haiti, where estimates of malaria seroprevalence varied substantially depending on the cutoff applied^16^. Cross-reactivity further complicates interpretation, particularly in regions with diverse immunological backgrounds, such as in African SARS-CoV-2 serosurveys, where unexpectedly high positivity rates were attributed to background reactivity from endemic infections ^22,23^. Similar issues arise in elimination contexts, where antibody persistence blurs the distinction between recent and past exposure; for example, Surendra et al. (2019) showed that conventional thresholds failed to separate long-term immunity from recent malaria infections in Indonesia^17^. Finally, the lack of well-characterized reference sera, a common problem in wildlife and zoonotic disease research, has been shown to inflate false-positive rates and limit assay validation^24^.

These difficulties underscore the need for analytical strategies that go beyond rigid thresholds. By combining Gaussian and skew-normal mixture models to accommodate asymmetry, applying rigorous goodness-of-fit testing to avoid spurious solutions, introducing a parsimonious score to ensure cluster stability, and leveraging hierarchical clustering to collapse weakly supported components into biologically meaningful groups, the decisional FMM framework directly addresses these barriers. Validation across CHIKV, SARS-CoV-2, and dengue datasets illustrates its broad relevance: in CHIKV, it produced nearly identical estimates to ROC-based cutoffs while identifying borderline discordant cases; in SARS-CoV-2, it captured disease heterogeneity and improved sensitivity without compromising specificity; and in dengue, it probabilistically distinguished exposed from unexposed children despite limited confirmed cases. Together, these findings highlight how the framework provides a reproducible and biologically grounded pathway for interpreting complex serological data across diverse epidemiological settings.

## Data Availability

All data produced in the present study are available upon reasonable request to the authors

